# A Learning Health System Approach to the COVID-19 Pandemic: System-Wide Changes in Clinical Practice and 30-Day Mortality Among Hospitalized Patients

**DOI:** 10.1101/2021.08.26.21262661

**Authors:** Erin K. McCreary, Kevin E. Kip, J. Ryan Bariola, Mark Schmidhofer, Tami Minnier, Katelyn Mayak, Debbie Albin, Jessica Daley, Kelsey Linstrum, Erik Hernandez, Rachel Sackrowitz, Kailey Hughes, Christopher Horvat, Graham M. Snyder, Bryan J. McVerry, Donald M. Yealy, David T. Huang, Derek C. Angus, Oscar C. Marroquin

## Abstract

**Introduction:** Rapid, continuous implementation of credible scientific findings and regulatory approvals is often slow in large, diverse health systems. The COVID-19 pandemic created a new threat to this common “slow to learn and adapt” model in healthcare. We describe how UPMC committed to a rapid learning health system (LHS) model to respond to the COVID-19 pandemic.

**Methods:** An observational cohort study was conducted among 11,429 hospitalized patients from 22 hospitals (PA, NY) with a primary diagnosis of COVID-19 infection (March 19, 2020 – June 6, 2021). Sociodemographic and clinical data were captured from UPMC electronic medical record (EMR) systems. Patients were grouped into four time-defined patient “waves” based on nadir of daily hospital admissions, with wave 3 (September 20, 2020 – March 10, 2021) split at its zenith due to high volume with steep acceleration and deceleration. Outcomes included changes in clinical practice (e.g., use of corticosteroids, antivirals, and other therapies) in relation to timing of internal system analyses, scientific publications, and regulatory approvals, along with 30-day rate of mortality over time.

**Results:** Mean (SD) daily number of hospital admissions was 26 (28) with a maximum 7-day moving average of 107 patients. System-wide implementation of the use of dexamethasone, remdesivir, and tocilizumab occurred within days of release of corresponding seminal publications and regulatory actions. After adjustment for differences in patient clinical profiles over time, each month of hospital admission was associated with an estimated 5% lower odds of 30-day mortality (adjusted OR = 0.95, 95% confidence interval: 0.92 – 0.97, *p* < .001).

**Conclusions:** In our large LHS, near real-time changes in clinical management of COVID-19 patients happened promptly as scientific publications and regulatory approvals occurred throughout the pandemic. Alongside these changes, patients with COVID-19 experienced lower adjusted 30-day mortality following hospital admission over time.

## INTRODUCTION

Integration of evidence-based practices is notoriously slow, especially at larger, diverse, health care systems. The emergence of a rapidly-spreading, severe respiratory virus pandemic created a heightened need for change in this common approach.^1,2^ Current research infrastructure and information technology systems facilitate unprecedented volume and speed of pandemic-related information, and data sharing in the biomedical literature, social media, and other resources allow insights to flow much more quickly.^3^ Making efficient, optimal use of this massive, constantly changing information is paramount to minimize the deadly impact of the pandemic, and for the health and welfare of humanity at large.

In addition to the need for coordinated global approaches to pandemics,^1^ individual health care delivery systems must seek to give equitable, evidence-based care across institutions regardless of geographical region or hospital type.^4^ In this realm, a *learning health system* (LHS) is an ideal organizing principle to inform evidence-based responses to public health emergencies like COVID-19.^4^ The LHS concept is characterized as an environment in which “science, informatics, incentives, and culture are aligned for continuous improvement and innovation, with best practices seamlessly embedded in the delivery process and new knowledge captured as an integral by-product of the delivery experience.”^5^

Seeking to embrace the LHS model, the UPMC health system leveraged its science, data, and analytics capabilities and established the multidisciplinary COVID-19 Therapeutics Committee in early 2020. The purpose of this Committee was to evaluate any possible COVID-19 treatment option and rapidly disseminate updated guidelines to all institutions within the system. The Committee also coordinated with information technology specialists to build forcing functions into several electronic medical records (EMRs) to enforce practice guideline recommendations, and also collaborated with research teams to integrate clinical practice with clinical trial enrollment across the enterprise. This LHS process, coupled with regular internal COVID-19 analyses from the UPMC Clinical Analytics Team (described in Methods), formed the basis for establishing, disseminating, and documenting data-driven clinical recommendations to all UPMC outpatient and in-patient facilities caring for patients with COVID-19.

We describe the UPMC LHS approach to the COVID-19 pandemic since March 2020. We share processes on the development and dissemination of clinical guidelines that occurred in a near real-time manner across the entire UPMC system. We also share quantitative results of how such changes mirrored credible findings and information from key scientific publications and regulatory approvals. This is followed by temporal assessment of the 30-day rate of mortality of hospitalized patients with COVID-19.

## QUESTION OF INTEREST

How have hospital patient volumes, patient clinical management, and 30-day mortality changed since the onset of the COVID-19 pandemic within a large, multi-hospital learning health system (LHS)?

## METHODS

### Sources of Data

We used data captured in the EMR and ancillary clinical systems, all of which are aggregated and harmonized in a Clinical Data Warehouse (CDW). UPMC is a 40-hospital integrated academic healthcare system providing care principally within central and western Pennsylvania (USA). For the 22 hospitals with complete EMR data in the CDW, we accessed all key clinical data, including detailed sociodemographic and medical history data, diagnostic and clinical tests conducted, surgical and other treatment procedures performed, prescriptions ordered, and billing charges on all outpatient and in-hospital encounters, with diagnoses and procedures coded based on the International Classification of Diseases, Ninth and Tenth revisions (ICD-9 and ICD-10, respectively).

### Influence of UPMC COVID-19 Therapeutics Committee

We evaluated the influence of the UPMC COVID-19 Therapeutics Committee on change in COVID-19 clinical practice by time series plotting of the prevalence of in-hospital use of selected medications in relation to internal analyses and key scientific publications and regulatory approvals routinely reviewed by the committee. The UPMC COVID-19 Therapeutics Committee charge was to continuously evaluate evidence to create and disseminate treatment recommendations across the UPMC system. The process included: (1) weekly to bi-weekly review of internal analyses of COVID-19 patient testing, clinical practice, and outcomes generated from the CDW; (2) interim review of results from UPMC-led Randomized Embedded Multifactorial Adaptive Platform for COVID-19 (REMAP-COVID) trials, a global adaptive platform for trials of hospitalized and ambulatory patients with COVID-19; ^6,7^ (3) weekly and ad-hoc review of key external scientific publications, press releases, and regulatory approvals of COVID-19 treatment approaches; (4) consensus determination of patient criteria and clinical instructions for use (and non-use) of established and emerging treatment approaches; (5) creation of EMR-embedded forcing functions to enforce therapeutics recommendations and guide prescribing at the point of care; (6) empowerment of local pharmacists to review and approve all COVID-19-related medications within the context of the guidelines; and (7) system-wide dissemination of continuously updated treatment recommendations using multimodal media sources.

The system-wide dissemination of treatment guidelines to all physicians and other clinicians affiliated with UPMC occurred through email notifications, computer screensavers, educational webinars, and formal directives from the chair of the Committee. A COVID-19 therapeutics webpage was built into the system intranet. The COVID-19 Therapeutics Committee also created continuous, updated recommendations on the use of monoclonal antibodies for ambulatory COVID-19 patients beginning in November 2020; however, the present analysis is restricted to treatment of hospitalized patients and omits that intervention.

### Patient Population

We identified 323,101 patients with nucleic acid amplification tests for SARS-CoV-2 performed at a UPMC facility during the period March 17, 2020 to June 6, 2021. Of 53,183 patients (16.5%) testing positive, 9,554 (18.0%) were hospitalized at one of 22 UPMC hospitals. An additional 1,875 COVID-19 patients were hospitalized at a UPMC hospital with testing performed outside the UPMC system, resulting in a total of 11,429 hospitalized patients for analysis (**Supplemental Figure S1**)

### Outcomes

We assessed changes in utilization of COVID-19 pharmacotherapy, level of oxygen support during hospitalization, and 30-day mortality from the index date of hospital admission. Pharmacotherapy and oxygen support were determined by the presence of charge codes within UPMC billing software. We assessed 30-day mortality by the hospital discharge disposition of “Ceased to Breathe” sourced from the inpatient Medical Record System, as well as deaths after discharge identified with the Death Master File (DMF) from the Social Security Administration (SSA)(NTIS 2021) as an external data source.

### Explanatory Variables

For assessment of temporal changes, we categorized the study analysis period into 4 discrete “waves” based on empirical change in hospital admissions within the UPMC system. We chose the 4-wave classification scheme (**Figure 1**) based on the start and nadir of individual waves. However, because Wave 3 (September 29, 2020 – March 10, 2021) had dramatically higher hospital admissions and discharges, we split this wave at its zenith to assess its impact during rapid acceleration and deceleration. For assessment of variation between waves, we considered demographic variables, clinical history and medical comorbidities, laboratory values, vital signs, and medication use, with a focus on indicators of changing clinical practice such as use and timing of specific medications. We further assessed changes in COVID-19 clinical practice by the date of important scientific and regulatory events, as formally reviewed by the UPMC COVID-19 Therapeutics Committee. We also assessed potential variation in clinical practice across the 22 hospitals by classification as “large academic” (*n*=5), “large community” (*n*=8), or “small community” (*n*=9) (**see Supplemental Table 1**).

**Figure 1.**
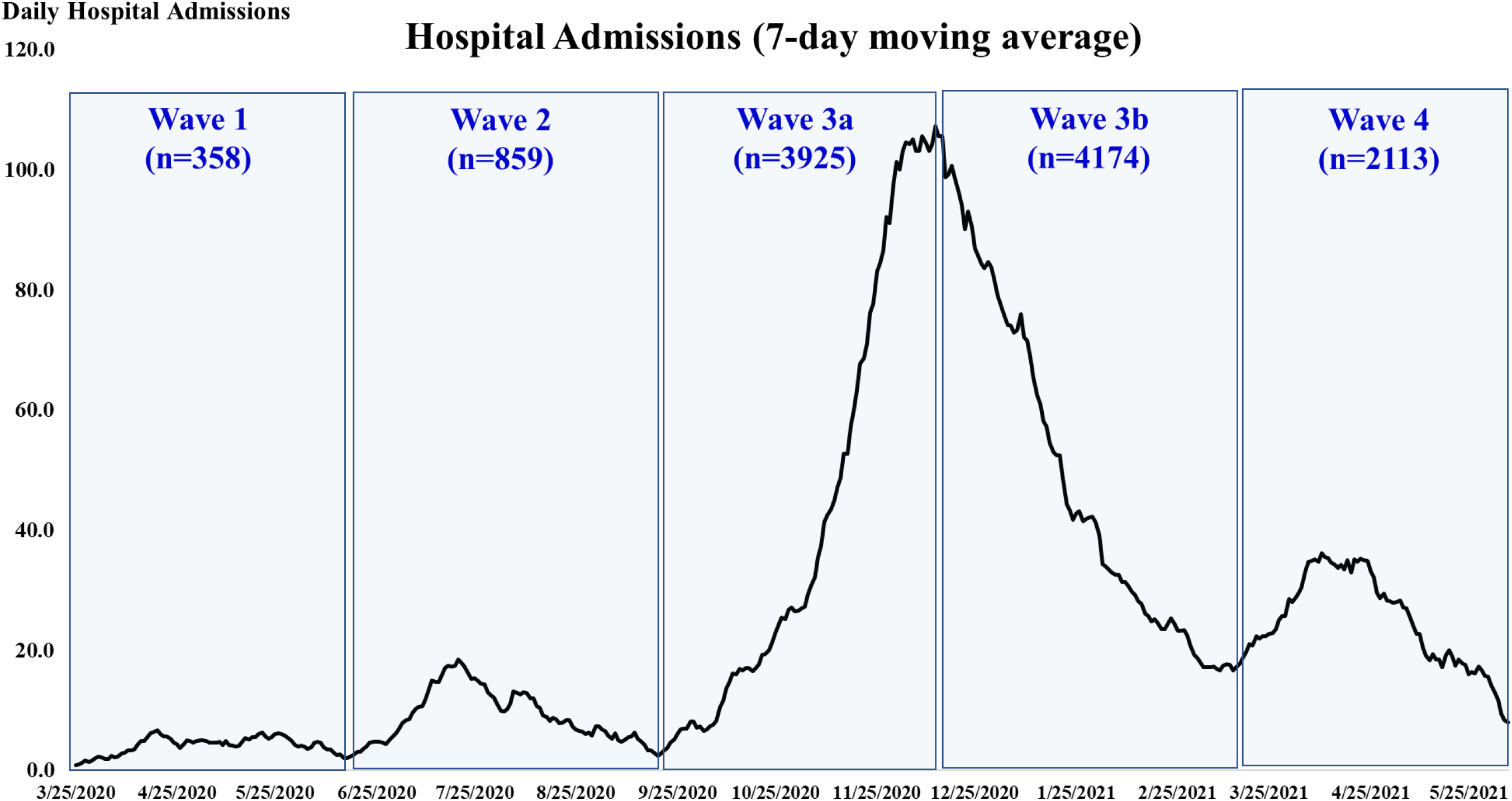
Plot of 7-day moving average of COVID-19 hospital admissions by empirically defined “waves” based on nadir and zenith (wave 3) of admissions. The time periods for the waves were as follows: Wave 1: - March 19 – June 16, 2020; Wave 2: June 17 – September 19, 2020; Wave 3a: September 20 - December 13, 2020; Wave 3b: December 14, 2020 – March 10, 2021; Wave 4: March 11 – June 6, 2021.

### Statistical Methods

We describe changes over time in COVID-19 hospital admissions using 7-day moving mean and median values. We plotted temporal changes in pharmacotherapy used in-hospital on a weekly basis and anchored to important scientific and regulatory events. Medication use and oxygen support (proportion of patients) plots by wave of hospital admission were evaluated by the Cochran-Mantel-Haenszel test of trend. We compared presenting characteristics of hospitalized patients across the 4 waves using analysis of variance (ANOVA) or Wilcoxon tests for continuous variables (based on distributional properties) and chi-square tests for categorical variables. Crude rates of 30-day mortality for test positive and hospitalized patients by wave were censored at May 7, 2021 (i.e. to allow 30-day follow-up for all patients). A logistic regression model was fit using 30-day mortality as the dependent variable with stepwise selection of pre-treatment explanatory variables. Date of hospital admission was added to the model at the last stage to assess whether the odds of 30-day mortality changed over time after adjustment for different patient characteristics. We did not impute missing values in any analyses. A two-sided type I error rate of 0.05 was used, and all analyses were conducted using the SAS System, Version 9.4 (SAS Institute, Cary, NC). We used The REporting of studies Conducted using Observational Routinely-collected health Data (RECORD) approach^8^ (see **Supplemental Table 2)**. Our study received formal ethics approval by the UPMC Ethics and Quality Improvement Review Committee (Project ID Project ID 2882).

## RESULTS

### Hospital Admissions

Over the 14+ month study period, the mean (SD) daily number of hospital admissions was 26 (28) with median of 17, IQR of 6 – 31, maximum 7-day moving mean of 107, and steep acceleration and deceleration during wave 3 from late September 2020 to early March 2021 (**Figure 1**). The mean hospital admission rate per day by wave was 4.0 (wave 1), 9.1 (wave 2), 46.1 (wave 3a), 48.0 (wave 3b), and 24.0 (wave 4).

### Temporal Changes in Clinical Practice

Among patients who received any form of supplemental oxygen, there was rapid system-wide implementation in the use of dexamethasone immediately around the date in which initial positive results of the RECOVERY trial were published as a pre-print,^9^ (**Figure 2**). Of note, subsequent peer-review publication^10^ did not trigger an added uptake in use of dexamethasone. A steep increase in the use of remdesivir among patients on oxygen therapy occurred after Emergency Use Authorization (EUA) granted by the FDA^11^ and subsequent public announcements and regulatory actions^12-14^ (**Figure 3**). There was no appreciable variation in the use of dexamethasone or remdesivir by volume or type across the 22 UPMC hospitals (**Supplemental Figures S2 and S3**).

**Figure 2.**
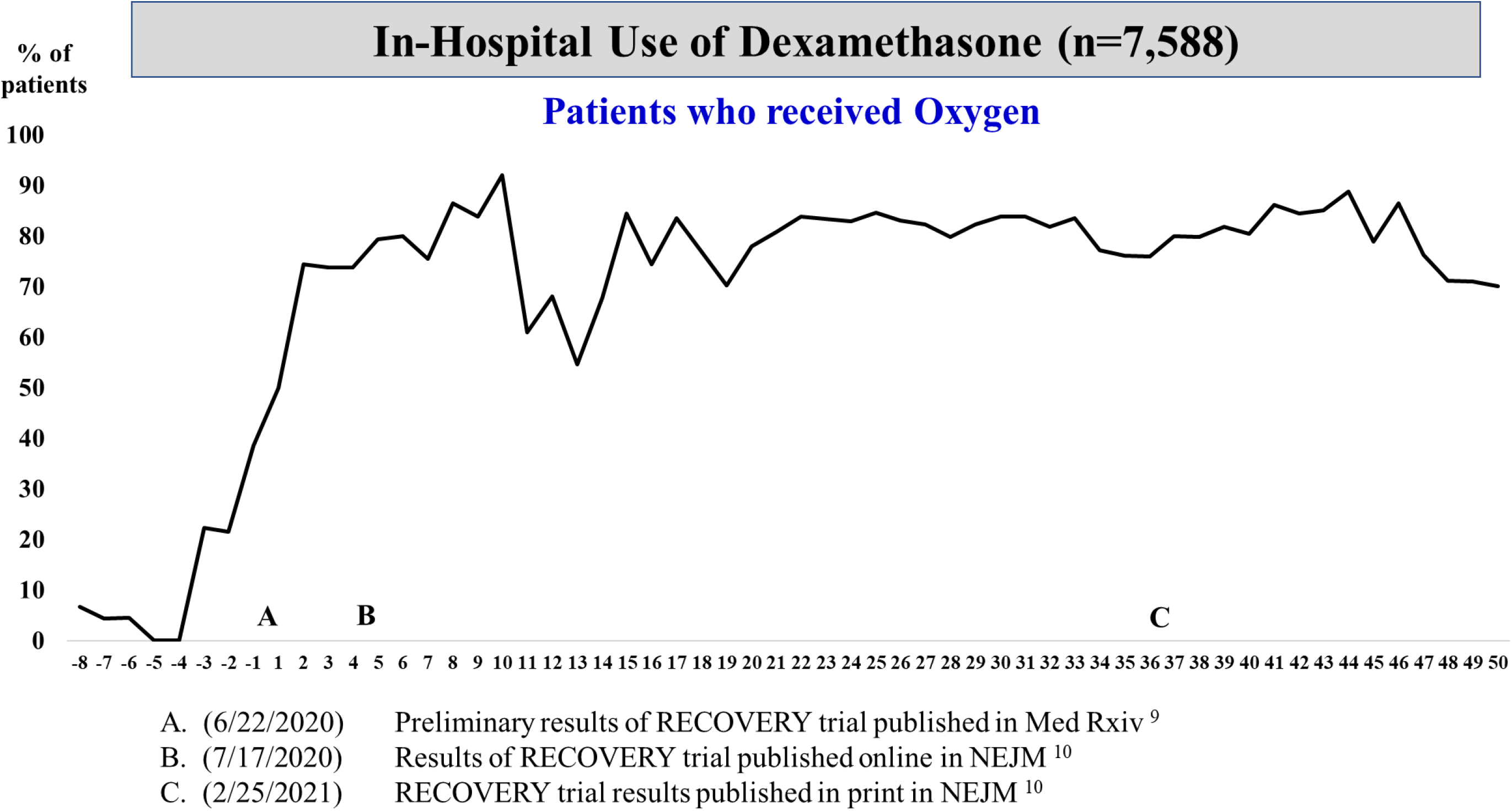
Plot of weekly prevalence (%) of in-hospital use of dexamethasone among patients who received oxygen. On the x-axis, negative numbers reflect weeks prior to seminal event “A”, the date (June 22, 2020) in which preliminary results of the RECOVERY trial were published in *Med Rxiv*. Positive numbers reflect weeks after seminal event A.

**Figure 3.**
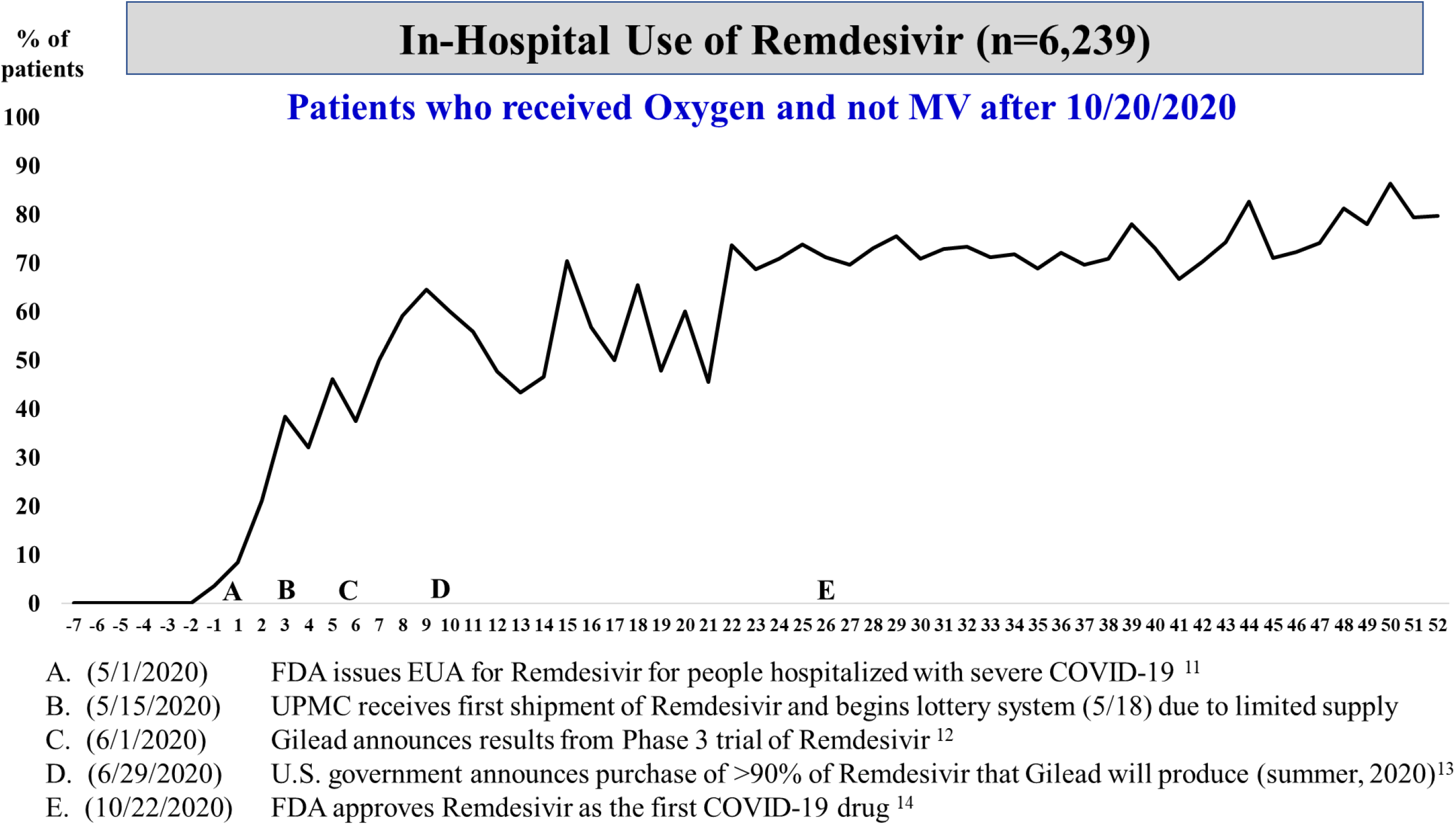
Plot of weekly prevalence (%) of in-hospital use of remdesivir among patients who received oxygen (but not mechanical ventilation after 10/20/2020). On the x-axis, negative numbers reflect weeks prior to seminal event “A”, the date (May 1, 2020) in which the FDA issued Emergency Use Authorization (EUA) for remdesivir for patients hospitalized with severe COVID-19.

In contrast, despite widespread publicity^15,16^ our group recommended no role for hydroxychloroquine outside of the context of a clinical trial. Subsequently, in-hospital use was very low (<6%) and did not vary over time, including after FDA EUA revocation of hydroxychloroquine,^17,18^ and thus may be represented solely by patients taking this medication for a non-COVID-19 indication or enrolled in a clinical trial (**Figure 4**). More recently, an increase in the use of tocilizumab among eligible patients occurred immediately following pre-print release of the REMAP-CAP trial results,^19^ (**Figure 5**) and a few weeks after publication of the RECOVERY^20^ and REMAP-CAP trial results^21^.

**Figure 4.**
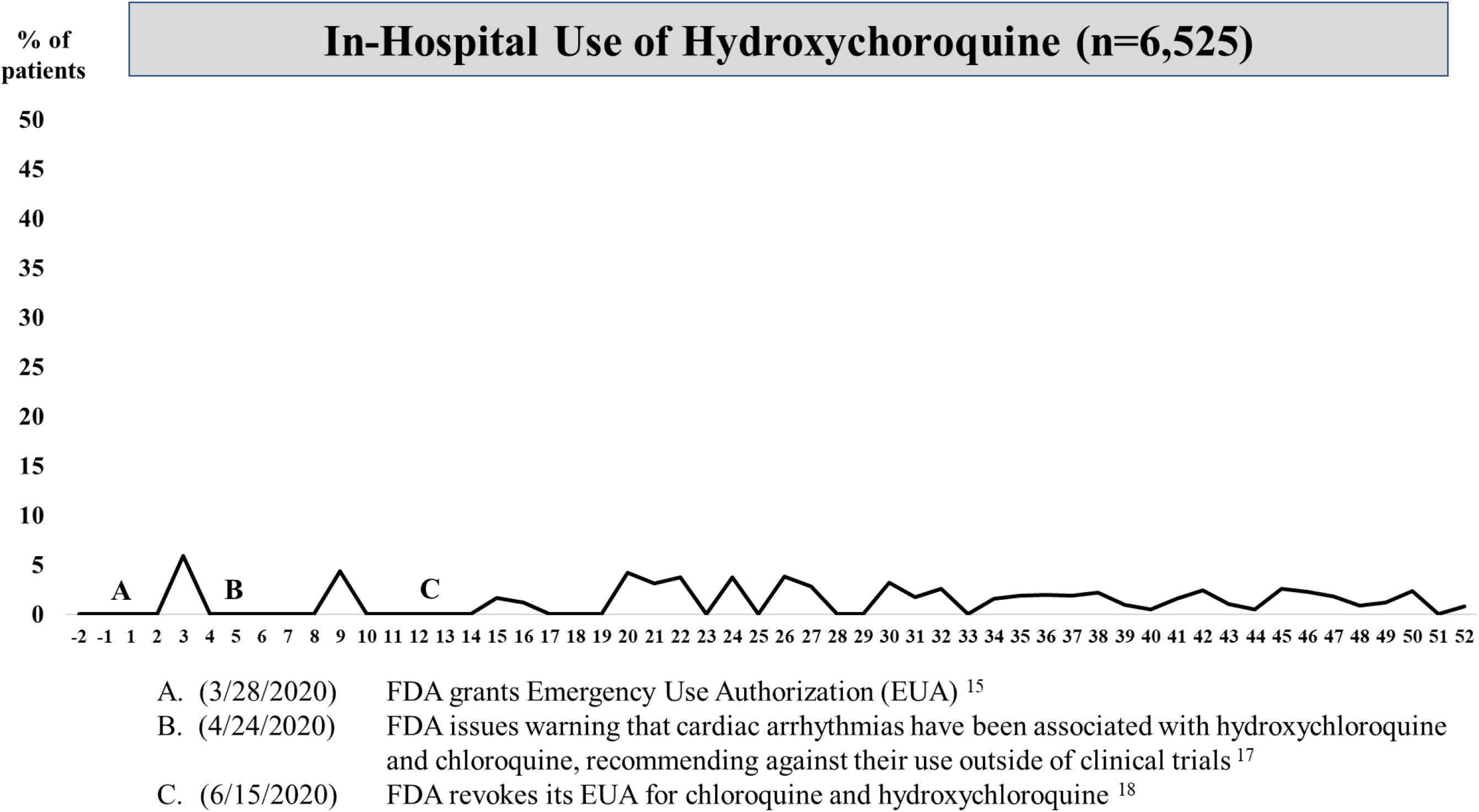
Plot of weekly prevalence (%) of in-hospital use of hydroxychloroquine. On the x-axis, negative numbers reflect weeks prior to seminal event “A”, the date (March 28, 2020) in which the FDA granted EUA of hydroxychloroquine for COVID-19 patients.

**Figure 5.**
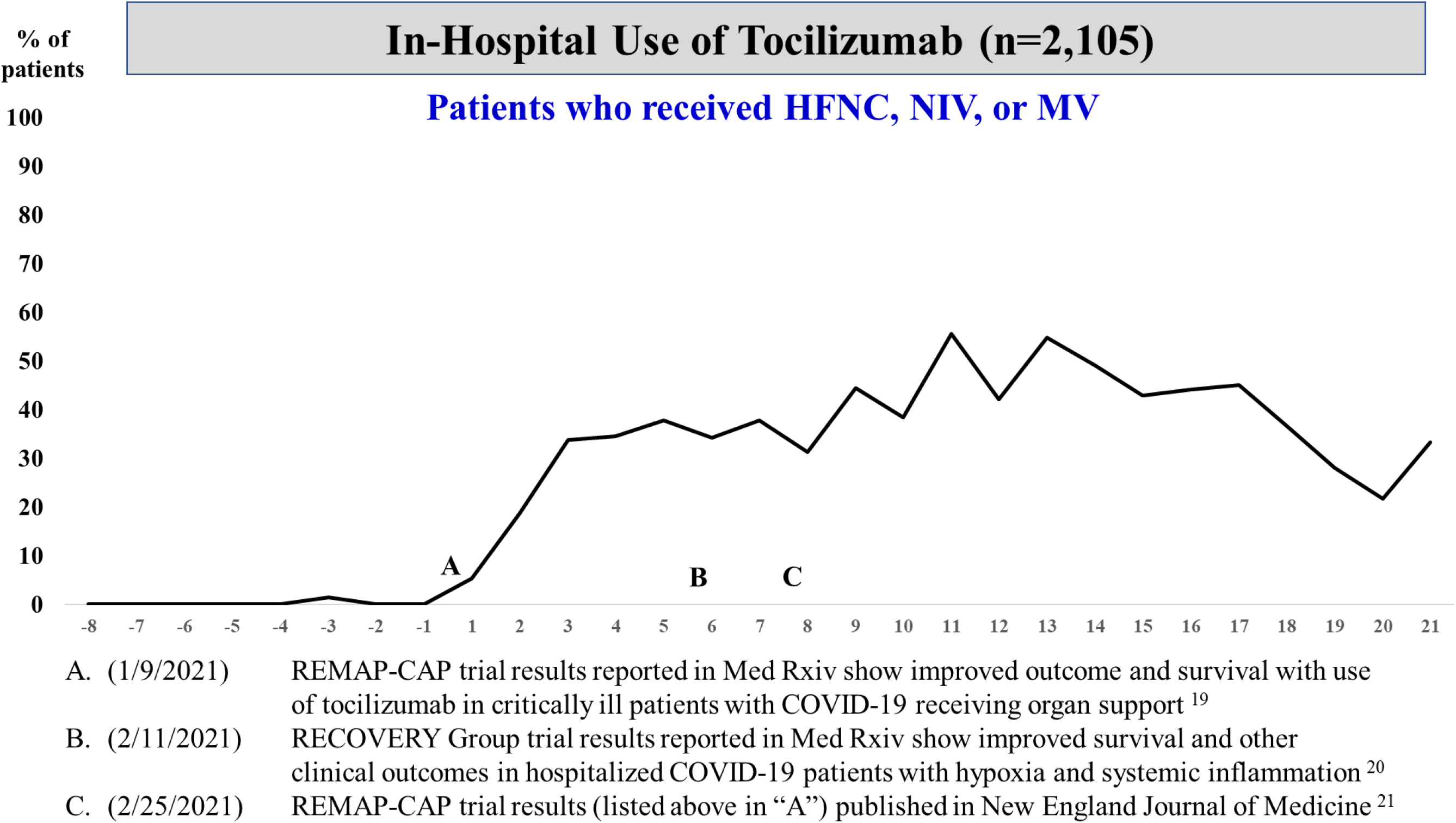
Plot of weekly prevalence (%) of in-hospital use of tocilizumab among patients who received high-flow nasal cannula (HFNC), BiPAP/CPAP (NIV), or mechanical ventilation (MV). On the x-axis, negative numbers reflect weeks prior to seminal event “A”, the date (January 9, 2021) in which tocilizumab trial results were published among critically ill patients with COVID-19 who were receiving organ support.

Among patients who received oxygen therapy, in-hospital use of corticosteroids was 80% or higher starting in wave 2. Two-thirds or more of all patients received steroids within 1 day of admission. Beginning in wave 3, about three-quarters of all clinically appropriate patients received remdesivir (almost always within 1 day of admission) (**Supplemental Figure S4**). Use of non-invasive ventilation did not vary appreciably across waves, whereas use of mechanical ventilation was markedly lower after wave 1 (**Supplemental Figure S4**).

### Temporal Changes in Patient Characteristics and 30-Day Mortality

Hospitalized patients in waves 1 and 3a/3b were significantly older than patients in wave 2 (about 3-4 years), and the most recent wave 4 patients were the youngest with mean (median) age of 59.5 (62) years and about a quarter (27%) being age 50 or younger (**Supplemental Table 3**). In aggregate, patients in waves 1 and 3 generally presented with more comorbidities, higher estimated 90-day probability of mortality, and higher neutrophil to lymphocyte ratio (NLR) and systemic inflammatory index than patients in waves 2 and 4 (**Supplemental Table 3**).

Among all test positive patients (hospitalized and not hospitalized), the 30-day mortality rate ranged from a high of 5.6% in wave 1 to a low of 2.2% in wave 4 (**Table 1**). Similar in direction, among hospitalized patients, the 30-day mortality rate ranged from a high of 19.8% in wave 1 to a low of 8.5% in wave 4. Consistent with different risk profiles across the 4 waves, 30-day mortality was highest in wave 1, intermediate in wave 3, and lowest in waves 2 and 4. After statistical adjustment, factors independently associated with 30-day mortality rate after hospitalization included older age (15% increased odds per 5 years), male gender (27% increased odds), estimated risk of mortality within 90 days after being hospitalized (12% increased odds per 5 percentage points), and higher white blood cell, ALT, and NLRs (**Table 2**). Of note, adjusting for different risk profiles, each month of hospital admission to the UPMC system was associated with an estimated 5% lower odds of 30-day mortality (adjusted OR = 0.95, 95% confidence interval: 0.92 – 0.97, *p* < .001).

**Table 1.** Thirty-Day Mortality Rate of Test Positive and Hospitalized Cases by Wave.

| **Wave** | **Patient Time Period** | **All Test Positives** |  | **Hospitalized Cases** |  |
| --- | --- | --- | --- | --- | --- |
|  |  | **N** | **Rate (%)** | **N** | **Rate (%)** |
| 1 | March 19 – June 16, 2020 | 1369 | 5.6 | 358 | 19.8 |
| 2 | June 17 – September 19, 2020 | 3926 | 2.5 | 859 | 10.1 |
| 3a | September 20 – December 13, 2020 | 21471 | 2.8 | 3925 | 14.9 |
| 3b | December 14, 2020 – March 10, 2021 | 18637 | 2.9 | 4174 | 13.0 |
| 4 | March 11 – May 7, 2021 | 6378 | 2.2 | 1674 | 8.5 |

**Table 2.** Logistic Regression Analysis of Factors Associated with 30-Day Mortality (n=10,763 Hospitalized Patients)

| **Factor** | **Unadjusted**<br>**OR** | **Adjusted**<br>**OR** | **95%**<br>**CI** | **p-**<br>**value** |
| --- | --- | --- | --- | --- |
| Age (per 5 years) | 1.23 | 1.15 | 1.11 – 1.18 | <.001 |
| Male gender | 1.41 | 1.27 | 1.12 – 1.43 | <.001 |
| Estimated risk of mortality within 90-days after<br>being hospitalized (per 5 percentage points) <sup>a</sup> | 1.17 | 1.12 | 1.10 – 1.14 | <.001 |
| Log white blood cell count at hospital admission | 1.98 | 1.52 | 1.36 – 1.70 | <.001 |
| Log alanine aminotransferase (ALT) at hospital<br>admission | 1.25 | 1.42 | 1.30 – 1.54 | <.001 |
| Neutrophil to lymphocyte ratio (per quintile) | 1.24 | 1.19 | 1.15 – 1.24 | <.001 |
| **Date of hospital admission (per month)** | **0.95** | **0.95** | **0.92 – 0.97** | **<.001** |

Model c statistic = 0.751 based on 1,416 deaths. <sup>a</sup> Risk score is derived from an internally validated algorithm that is comprised of a range of variables predictive of mortality including socio-demographics, medical history, recent laboratory values, and prior health care utilization.

## DISCUSSION

In 2009, the National Academy of Medicine (NAM) called for development of a learning health system (LHS), setting a goal that by 2020, “…90 percent of clinical decisions will be supported by accurate, timely, and up-to-date clinical information, and will reflect the best available evidence.”^22^ The importance of this LHS goal is emblematic with the COVID-19 pandemic.^23^ While lacking the ability to demonstrate cause and effect, the fact that the adjusted risk of in-hospital mortality among hospitalized COVID-19 patients at UPMC hospitals has decreased monthly by an average of 5% suggests a consistent learning effect to improved patient care. Additionally, there was no appreciable variation in type or volume of pharmacotherapy agents utilized for patients with COVID-19 across 22 hospitals, achieving the goal of equity and access regardless of patient zip code. Importantly, this model continues and can rapidly adapt as needed for SARS-CoV-2 variants, vaccination efforts, and other key variables.

In addition to continuous evaluation of UPMC internal analyses and controlled clinical trials, the COVID-19 Therapeutics Committee has evaluated the surge of COVID-19-related preprints and peer-reviewed publications that have emanated on an unprecedented scale throughout the pandemic.^24,25,26^ This placed a premium on expertise in evaluating the merits of published information. While our committee recognized the benefits of and thus implemented steroids, remdesivir, and tocilizumab in selected COVID-19 patients, it refuted use of hydroxychloroquine despite its EUA, given the existing data.^27^

The time between clinical evidence arising and uniform implementation of use was in days-to-weeks, rather than months-to-years, which has been the traditional gap for implementation of findings from RCTs into clinical practice.^28,29,30^ We invested substantial efforts in the use of near-real time data and evidence (as per NAM LHS guidance), especially when the lack of available therapies fueled adoption of both warranted and unwarranted treatments. The average monthly risk-adjusted decrease in mortality of 5% observed in our healthcare system is noteworthy given the overall worse clinical profile of patients seen in wave 3. While utilization of pharmacotherapy is the focus of this analysis, it is likely that the observed improvements are multifactorial in nature. Alongside the Therapeutics Committee, an intensive care unit management group made real-time recommendations surrounding respiratory support strategies and other critical, supportive care and a systemwide infection prevention taskforce guided testing, tracing, isolation, and use of personal protective equipment. Accordingly, we posit there were changes of unmeasured practice patterns (i.e., ventilation strategies) learned over time that also contributed to the improved outcomes, and the rapid implementation of approved pharmacotherapies is a surrogate marker of systemwide learning. Lastly, while improvements in outcomes over time are natural to the progression of science and medical practice, the fact that the improvement seen in our healthcare system happened in a short time and mostly prior to mass vaccination, speaks, at least in part, to the importance of our system’s embracing the organizational push to be an LHS.

While desirable, no formal criteria or certification process exist for an institution to be designated as an LHS.^4^ One component we believe is essential is embedding of randomized controlled trial procedures into routine care processes using existing institutional infrastructure and electronic health records.^6^ This approach defines broad eligibility criteria and aims to enroll as many “real-world” patients as possible to continuously evaluate therapies believed to be potentially efficacious. The key is avoiding “research” and “care” schisms, but rather use all care as an opportunity to learn about care improvement. Randomization is an added tool for some efforts, allowing adaptation as the trial evolves such that subjects are preferentially randomized to receive better performing arms based on interim analyses—termed “response adaptive randomization.”^31^ This was accomplished at our hospitals by embedding REMAP-CAP enrollment into the EMR, screening all patients with COVID-19 at all hospitals for trial eligibility, and integrating trial enrollment with Therapeutics Committee treatment guidelines.^32^

There are some limitations to our study; because this is the experience in one, albeit large, integrated healthcare system in Western Pennsylvania, the generalizability of our findings may be questioned. However, the fact that we saw similar findings across our different sites suggests that our findings are applicable across academic, community, and rural hospitals. In addition, we cannot determine the extent to which the therapeutic interventions implemented uniformly by the UPMC COVID-19 Therapeutics Committee contributed to lower adjusted mortality over time, as opposed to other less well documented clinical practices that may have been implemented over time (i.e., mechanical ventilation).

The LHS description and results presented herein are not meant to be content- or institution-specific, but rather to illustrate some of the processes that can be used to support the NAM imperative for clinical decisions that are supported by accurate, timely, and up-to-date clinical information that reflects the best available evidence.^22^ On a broader level, we support the stated advocacy for a learning health network that promotes collaboration among health systems, community-based organizations, and government agencies, especially during public health emergencies.^4^

## CONCLUSION

Other institutions have qualitatively described their respective LHS processes employed in response to the COVID-19 pandemic,^33,34^ with limited quantitative temporal assessment of clinical outcomes.^35^ We believe our analysis and description is the first to empirically document how COVID-specific processes employed within an LHS were actually implemented to achieve timely changes in clinical practice on a system level, which likely in turn favorably influenced clinical outcomes as evidenced by lower mortality over time. We recommend that institutions in describing their respective LHS do so by linking (and presenting) processes and sources of information that were used in the establishment and dissemination of clinical care guidelines with data-documented temporal changes in clinical practice and patient outcomes.

## Data Availability

The data used to compile this manuscript are proprietary and confidential. The corresponding author will consider all research requests for subsequent analyses related to these data.

## Acknowledgement

The authors thank the administrative and clinical staff of the UPMC COVID-19 Therapeutics Committee, including but not limited to: Allison Hydzik, Larry Hruska, Jennifer Dueweke, Robert Shulik, Amy Lukanski, Rozalyn Russell, Debra Rogers, Jesse Duff, Kevin Pruznak, Jennifer Zabala, Trudy Bloomquist, Daniel Gessel, LuAnn King, Jonya Brooks, Libby Shumaker, Betsy Tedesco, Sarah Sakaluk, Kathleen Flinn, Susan Spencer, Le Ann Kaltenbaugh, Michelle Adam, Meredith Axe, Melanie Pierce, Debra Masser, Theresa Murillo, Sherry Casali, Jim Krosse, Jeana Colella, Rebecca Medva, Jessica Fesz, Ashley Beyerl, Jodi Ayers, Hilary Maskiewicz, Mikaela Bortot, Amy Helmuth, Heather Schaeffer, Janice Dunsavage, Erik Hernandez, Ken Trimmer, Sheila Kruman, Teressa Polcha, Kevin Collins, Al L’Altrelli, Alex Viehman, Alyssa Lopus, Amy Dutko, Asmaa Debri, Brandon Smith, Brian Campfield, Bryan McVerry, Crystal Gilbert, Diana Pakstis, Elise Martin, Erin Weslander, Glenn Rapsinski, Janice Dunsavage, Jennifer Kozar, Jill Breton, John Goldman, Kailey Hughes, Kirk Jones, Krystina Zaradzki, Luis Urrutia, Marian Michaels, Matthew Partsch, Michael Donohoe, Minh-Hong Nguyen, Ricardo Arbulu Guerra, Stacey Miske, Thomas Hebert, Tina Khadem, Anthony Pizon, and their entire teams. We also thank the U.S. federal government and Pennsylvania Department of Health for the provision of remdesivir and monoclonal antibody treatment.

## Figure Legends

**Supplemental Figure S1.**
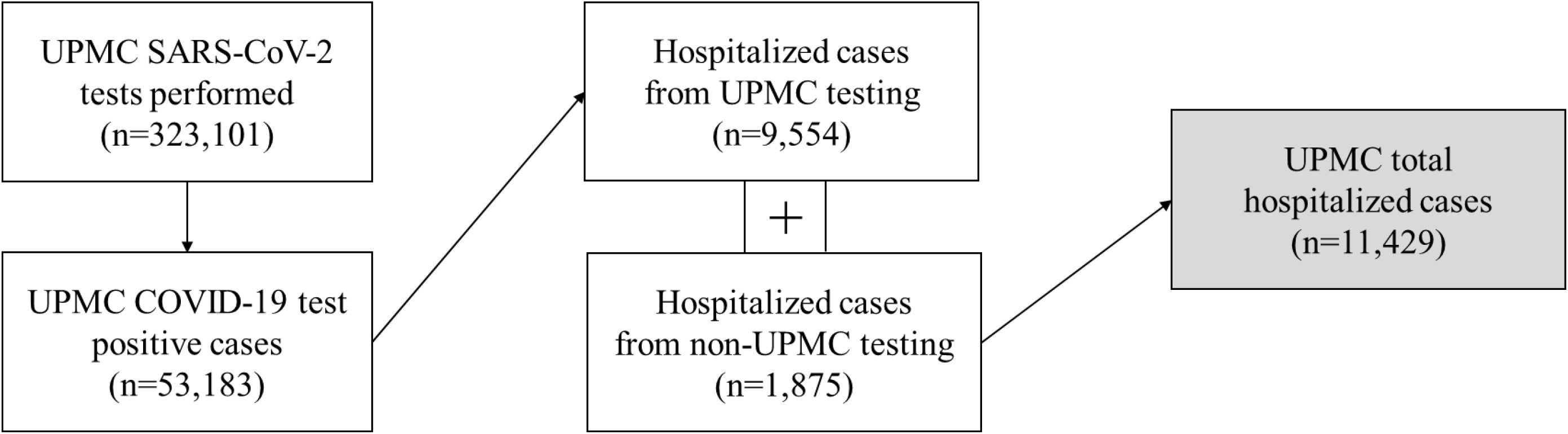
Diagram of SARS-CoV-2 testing performed within the UPMC system, including COVID-19 hospitalized patients aggregated from patients tested within and outside of the UPMC system.

**Supplemental Figure S2.**
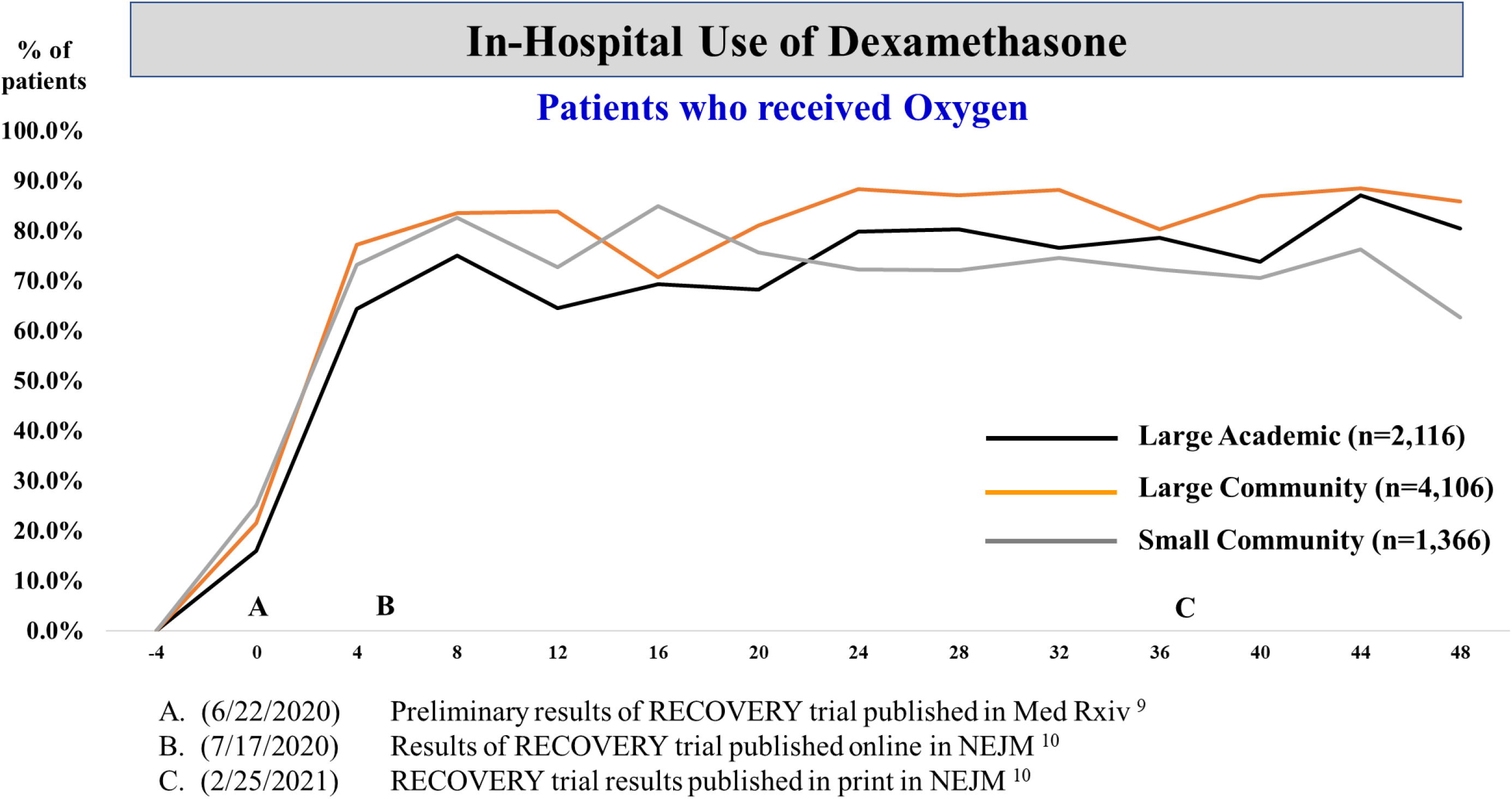
Plot of 4-week prevalence (%) of in-hospital use of dexamethasone among patients who received oxygen by hospital classification. On the x-axis, negative numbers reflect weeks prior to seminal event “A”, the date (June 22, 2020) in which preliminary results of the RECOVERY trial were published in *Med Rxiv*. Positive numbers reflect weeks after seminal event A.

**Supplemental Figure S3.**
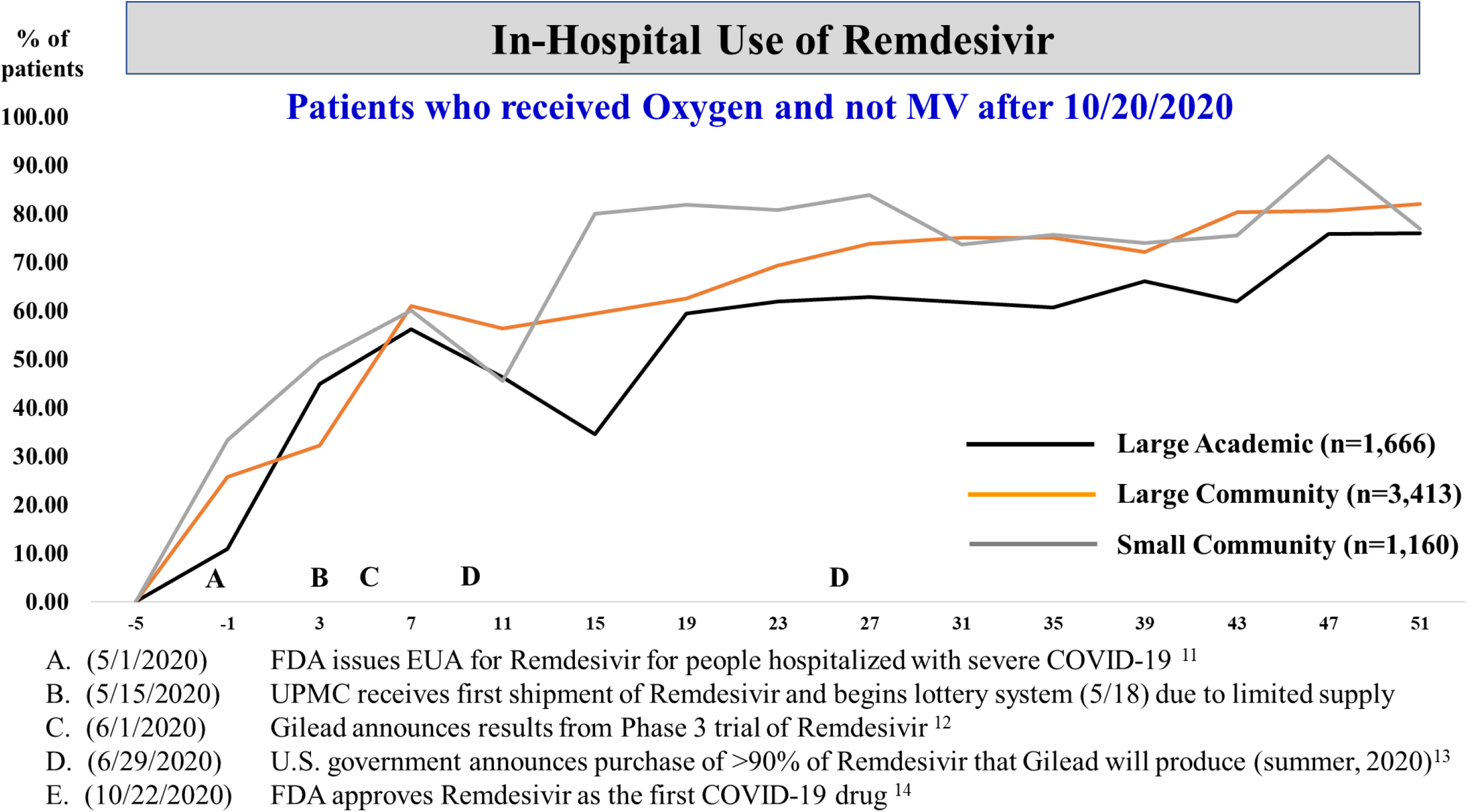
Plot of 4-week prevalence (%) of in-hospital use of remdesivir among patients who received oxygen (but not mechanical ventilation after 10/20/2020) by hospital classification. On the x-axis, negative numbers reflect weeks prior to seminal event “A”, the date (May 1, 2020) in which the FDA issued Emergency Use Authorization (EUA) for remdesivir for patients hospitalized with severe COVID-19.

**Supplemental Figure S4.**
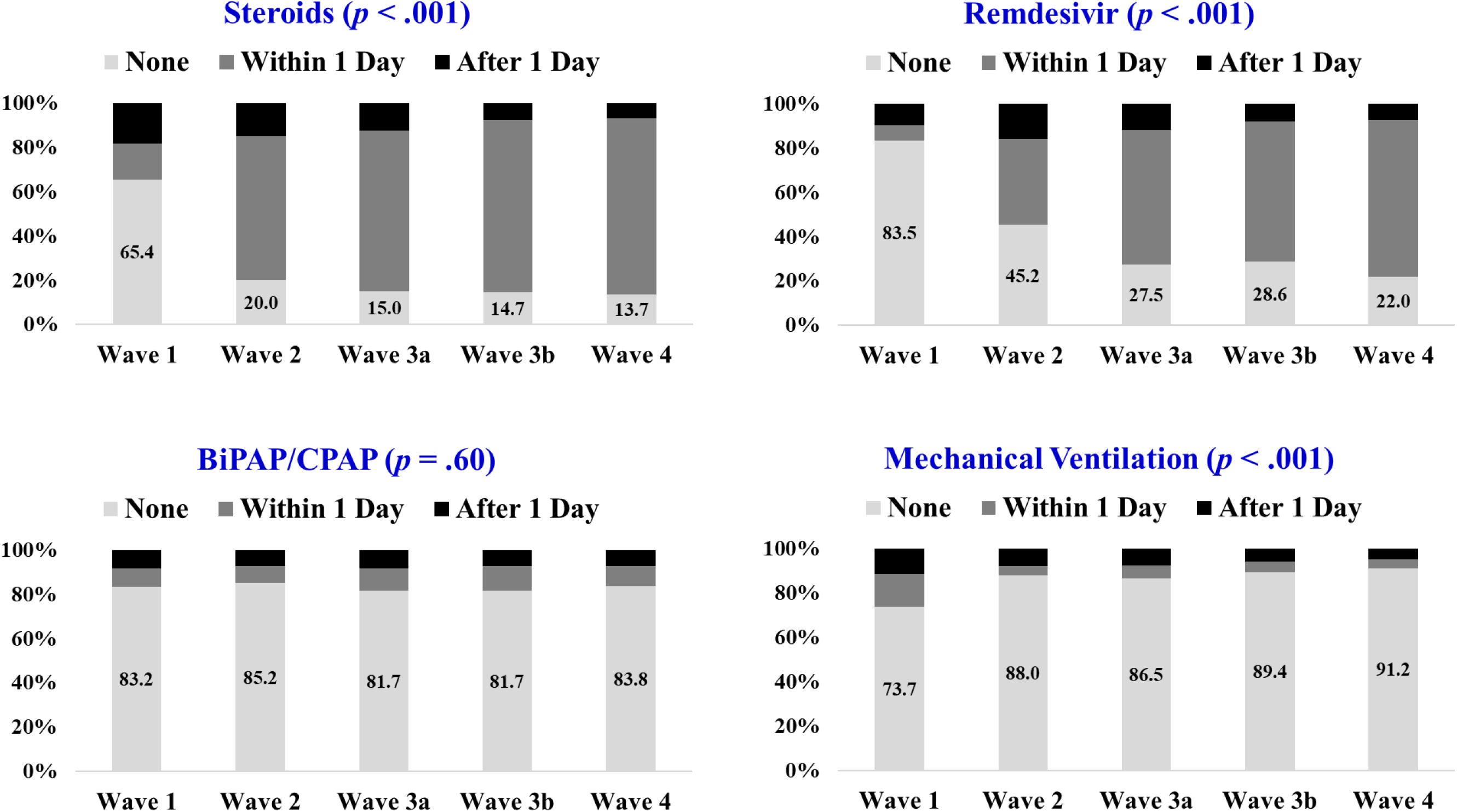
Stacked bar charts (100%) of the percentage of hospitalized patients treated with steroids (upper left), remdesivir (upper right), BiPAP/CPAP (lower left), and mechanical ventilation (lower right) by wave of hospital admission. For steroids, the denominator is patients on oxygen therapy. For Remdesivir, the denominator is patients on oxygen therapy and not mechanical ventilation after 10/20/2020. Light shading: treatment not used; intermediate shading: treated provided within 1 day of hospital admission; darker shading: treatment provided after the first day of hospital admission. P-values are based on the Cochran-Mantel-Haenszel test of trend.

---

**Supplemental Table 1.**
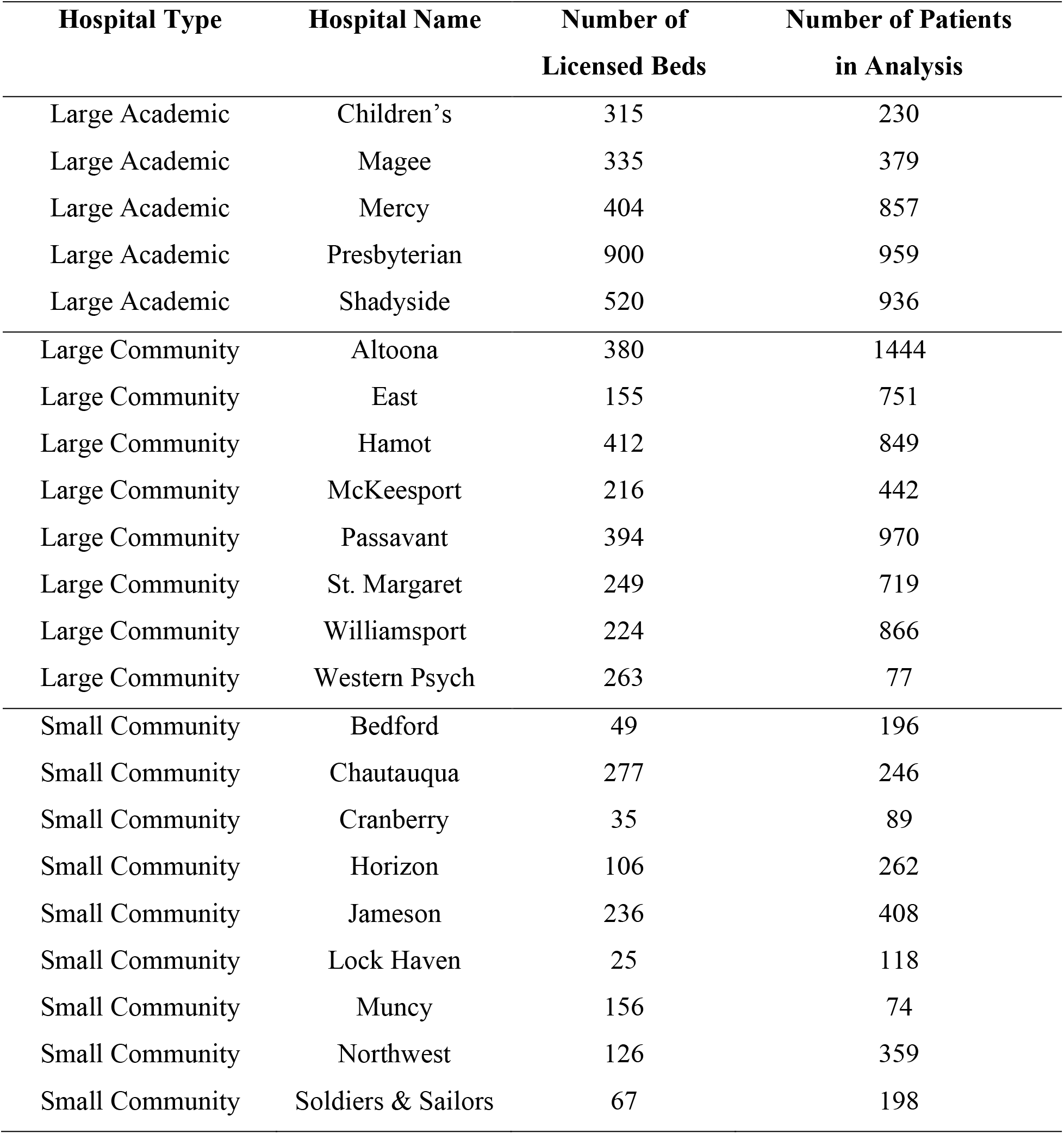
Listing of UPMC Hospitals by Type, Bed Capacity, and Volume.

| **Hospital Type** | **Hospital Name** | **Number of**<br>**Licensed Beds** | **Number of Patients**<br>**in Analysis** |
| --- | --- | --- | --- |
| Large Academic | Children's | 315 | 230 |
| Large Academic | Magee | 335 | 379 |
| Large Academic | Mercy | 404 | 857 |
| Large Academic | Presbyterian | 900 | 959 |
| Large Academic | Shadyside | 520 | 936 |
| Large Community | Altoona | 380 | 1444 |
| Large Community | East | 155 | 751 |
| Large Community | Hamot | 412 | 849 |
| Large Community | McKeesport | 216 | 442 |
| Large Community | Passavant | 394 | 970 |
| Large Community | St. Margaret | 249 | 719 |
| Large Community | Williamsport | 224 | 866 |
| Large Community | Western Psych | 263 | 77 |
| Small Community | Bedford | 49 | 196 |
| Small Community | Chautauqua | 277 | 246 |
| Small Community | Cranberry | 35 | 89 |
| Small Community | Horizon | 106 | 262 |
| Small Community | Jameson | 236 | 408 |
| Small Community | Lock Haven | 25 | 118 |
| Small Community | Muncy | 156 | 74 |
| Small Community | Northwest | 126 | 359 |
| Small Community | Soldiers & Sailors | 67 | 198 |

**Supplemental Table S2.** Checklist: The REporting of studies Conducted using Observational Routinely-Collected health Data (RECORD) statement.

|  | Item No. | STROBE items | Location in manuscript where items are reported | RECORD items | Location in manuscript where items are reported |
| --- | --- | --- | --- | --- | --- |
| **Title and abstract** |  |  |  |  |  |
|  | 1 | (a) Indicate the study's design with a commonly used term in the title or the abstract (b) Provide in the abstract an informative and balanced summary of what was done and what was found | Abstract, Pages 3-4 | RECORD 1.1: The type of data used should be specified in the title or abstract. When possible, the name of the databases used should be included.<br>RECORD 1.2: If applicable, the geographic region and timeframe within which the study took place should be reported in the title or abstract.<br>RECORD 1.3: If linkage between databases was conducted for the study, this should be clearly stated in the title or abstract. | Abstract, Page 3<br>Abstract, Page 3<br>Abstract, Page 3 |
| **Introduction** |  |  |  |  |  |
| Background rationale | 2 | Explain the scientific background and rationale for the investigation being reported | Introduction, Pages 5-6 |  |  |
| Objectives | 3 | State specific objectives, including any prespecified hypotheses | Introduction, Page 6 |  |  |
| **Methods** |  |  |  |  |  |
| Study Design | 4 | Present key elements of study design early in the paper | Methods, Pages 6-9 |  |  |
| Setting | 5 | Describe the setting, locations, and relevant dates, including periods of recruitment, exposure, follow-up, and data collection | Methods, Pages 6-9 |  |  |
| Participants | 6 | (a) *Cohort study* - Give the eligibility criteria, and the sources and methods of selection of participants. Describe methods of follow-up | Methods, Pages 8-9 | RECORD 6.1: The methods of study population selection (such as codes or algorithms used to identify subjects) should be listed in detail. If this is not possible, an explanation should be provided. | Methods, Pages 8-9 |
|  |  | *Case-control study* - Give the eligibility criteria, and the sources and methods of case ascertainment and control selection. Give the rationale for the choice of cases and controls<br>*Cross-sectional study* - Give the eligibility criteria, and the sources and methods of selection of participants<br>(b) *Cohort study* - For matched studies, give matching criteria and number of exposed and unexposed<br>*Case-control study* - For matched studies, give matching criteria and the number of controls per case | N/A | RECORD 6.2: Any validation studies of the codes or algorithms used to select the population should be referenced. If validation was conducted for this study and not published elsewhere, detailed methods and results should be provided.<br>RECORD 6.3: If the study involved linkage of databases, consider use of a flow diagram or other graphical display to demonstrate the data linkage process, including the number of individuals with linked data at each stage. | Methods, Pages 6-7, described in text |
| Variables | 7 | Clearly define all outcomes, exposures, predictors, potential confounders, and effect modifiers. Give diagnostic criteria, if applicable. | Methods, Pages 8-9 | RECORD 7.1: A complete list of codes and algorithms used to classify exposures, outcomes, confounders, and effect modifiers should be provided. If these cannot be reported, an explanation should be provided. | The coding of all variables across the different EMRs is too voluminous to individually report. |
| Data sources/<br>measurement | 8 | For each variable of interest, give sources of data and details of methods of assessment (measurement). Describe comparability of assessment methods if there is more than one group | Methods, Pages 8-9 |  |  |
| Bias | 9 | Describe any efforts to address potential sources of bias | Methods, Page 9 |  |  |
| Study size | 10 | Explain how the study size was arrived at | Methods, Pages 7-8 |  |  |
| Quantitative variables | 11 | Explain how quantitative variables were handled in the analyses. If applicable, describe which groupings were chosen, and why | Methods, Pages 8-9 |  |  |
| Statistical methods | 12 | (a) Describe all statistical methods, including those used to control for confounding<br>(b) Describe any methods used to examine subgroups and interactions<br>(c) Explain how missing data were addressed<br>(d) *Cohort study* - If applicable, explain how loss to follow-up was addressed<br>*Case-control study* - If applicable, explain how matching of cases and controls was addressed<br>*Cross-sectional study* - If applicable, describe analytical methods taking account of sampling strategy<br>(e) Describe any sensitivity analyses | Methods, Page 9<br>Methods, Page 9<br>Methods, Page 9-10<br>N/A |  |  |
| Data access and cleaning methods |  | .. |  | RECORD 12.1: Authors should describe the extent to which the investigators had access to the database population used to create the study population.<br>RECORD 12.2: Authors should provide information on the data cleaning methods used in the study. | Methods, Pages 7-9<br>Methods, Pages 7-9 |
| Linkage |  | .. |  | RECORD 12.3: State whether the study included person-level, institutional-level, or other data linkage across two or more databases. The methods of linkage and methods of linkage quality evaluation should be provided. | Methods, Pages 8-9 |
| **Results** |  |  |  |  |  |
| Participants | 13 | (a) Report the numbers of individuals at each stage of the study (*e.g.*, numbers potentially eligible, examined for eligibility, confirmed eligible, included in the | Methods, Pages 8-9 | RECORD 13.1: Describe in detail the selection of the persons included in the study (*i.e.*, study population selection) including filtering based on data quality, data availability and linkage. The | Methods, Pages 8-9<br>Supplemental Figure S1 |
|  |  | study, completing follow-up, and analysed)<br>(b) Give reasons for non-participation at each stage.<br>(c) Consider use of a flow diagram | Methods, Pages 8-9<br>Supplemental Figure S1 | selection of included persons can be described in the text and/or by means of the study flow diagram. |  |
| Descriptive data | 14 | (a) Give characteristics of study participants (*e.g.*, demographic, clinical, social) and information on exposures and potential confounders<br>(b) Indicate the number of participants with missing data for each variable of interest<br>(c) *Cohort study* - summarise follow-up time (*e.g.*, average and total amount) | Results, pages 10-11,<br>Supplemental Table 3<br>Not listed due to extensive number of variables examined<br>Results, Page 9 |  |  |
| Outcome data | 15 | *Cohort study* - Report numbers of outcome events or summary measures over time<br>*Case-control study* - Report numbers in each exposure category, or summary measures of exposure<br>*Cross-sectional study* - Report numbers of outcome events or summary measures | Results, Pages 11-12,<br>Table 1 |  |  |
| Main results | 16 | (a) Give unadjusted estimates and, if applicable, confounder-adjusted estimates and their precision (*e.g.*, 95% confidence interval). Make clear which confounders were adjusted for and why they were included<br>(b) Report category boundaries when continuous variables were categorized<br>(c) If relevant, consider translating estimates of relative risk into absolute risk for a meaningful time period | Results, Pages 11-12,<br>Tables 1 and 2<br>Results, Pages 9-11 |  |  |
| Other analyses | 17 | Report other analyses done—e.g., analyses of subgroups and interactions, and sensitivity analyses | Results, Page 10 |  |  |
| **Discussion** |  |  |  |  |  |
| Key results | 18 | Summarise key results with reference to study objectives | Discussion, page 12-13 |  |  |
| Limitations | 19 | Discuss limitations of the study, taking into account sources of potential bias or imprecision. Discuss both direction and magnitude of any potential bias | Discussion, Page 14 | RECORD 19.1: Discuss the implications of using data that were not created or collected to answer the specific research question(s). Include discussion of misclassification bias, unmeasured confounding, missing data, and changing eligibility over time, as they pertain to the study being reported. | Discussion, Page 14 |
| Interpretation | 20 | Give a cautious overall interpretation of results considering objectives, limitations, multiplicity of analyses, results from similar studies, and other relevant evidence | Discussion, Pages 12-14 |  |  |
| Generalisability | 21 | Discuss the generalisability (external validity) of the study results | Discussion, Page 14 |  |  |
| **Other Information** |  |  |  |  |  |
| Funding | 22 | Give the source of funding and the role of the funders for the present study and, if applicable, for the original study on which the present article is based | Title Page, Page 1 |  |  |
| Accessibility of protocol, raw data, and programming code |  | .. |  | RECORD 22.1: Authors should provide information on how to access any supplemental information such as the study protocol, raw data, or programming code. | Supplemental information including raw data are not permissible from this analysis. |

\*Reference: Benchimol EI, Smeeth L, Guttman A, Harron K, Moher D, Petersen I, Sørensen HT, von Elm E, Langan SM, the RECORD Working Committee. The REporting of studies Conducted using Observational Routinely-collected health Data (RECORD) Statement. *PLoS Medicine* 2015; in press.
\*Checklist is protected under Creative Commons Attribution ([CC BY](https://creativecommons.org/licenses/by/4.0/)) license.

**Supplemental Table 3.** Presenting Characteristics of Hospitalized Patients by Admission Wave.

| Characteristic | Wave 1 | Wave 2 | Wave 3a | Wave 3b | Wave 4 | P-value |
| --- | --- | --- | --- | --- | --- | --- |
|  | (N=358) | (N=859) | (N=3925) | (N=4174) | (N=2113) |  |
| Age in years, mean, median | 66.3, 69 | 64.3, 67 | 68.7, 72 | 67.7, 70 | 59.5, 62 | <.001 |
| Age 50 years or younger, (No.), % | (53) 14.8 | (179) 20.8 | (495) 12.6 | (579) 13.9 | (565) 26.7 | <.001 |
| Female gender, (No.), % | (184) 51.4 | (422) 49.1 | (1887) 48.1 | (2004) 48.0 | (1073) 50.8 | .19 |
| Race, (No.), % |  |  |  |  |  |  |
| White | (220) 63.6 | (575) 69.2 | (3284) 86.0 | (3590) 88.2 | (1581) 77.2 | <.001 |
| Black | (122) 35.3 | (233) 28.0 | (467) 12.2 | (433) 10.6 | (453) 22.1 |  |
| Other | (4) 1.2 | (23) 2.8 | (67) 1.8 | (49) 1.2 | (15) 0.7 |  |
| Smoking history, (No.), % |  |  |  |  |  |  |
| Current | (18) 9.6 | (41) 7.4 | (168) 6.3 | (187) 6.7 | (149) 11.3 | <.001 |
| Former | (74) 39.6 | (246) 44.2 | (1247) 46.8 | (1324) 47.4 | (538) 40.8 |  |
| Never | (95) 50.8 | (269) 48.4 | (1250) 46.9 | (1283) 45.9 | (632) 47.9 |  |
| Current alcohol use, (No.), % | (49) 27.2 | (185) 34.3 | (900) 34.6 | (920) 33.7 | (422) 33.5 | .36 |
| Current illicit drug use, (No.), % | (6) 3.4 | (21) 4.0 | (82) 3.2 | (83) 3.1 | (49) 4.0 | .60 |
| **Medical History** |  |  |  |  |  |  |
| History of diabetes, (No.), % | (71) 36.2 | (210) 37.3 | (998) 37.1 | (1053) 37.2 | (429) 32.0 | .01 |
| History of hypertension, (No.), % | (115) 58.7 | (345) 61.3 | (1788) 66.5 | (1852) 65.5 | (830) 61.9 | .004 |
| History of portal hypertension, (No.), % | (33) 16.8 | (72) 12.8 | (334) 12.4 | (332) 11.7 | (154) 11.5 | .24 |
| History of hyperlipidemia, (No.), % | (103) 52.6 | (311) 55.2 | (1605) 59.7 | (1667) 58.9 | (710) 53.0 | <.001 |
| History of morbid obesity, (No.), % | (49) 25.0 | (141) 25.0 | (609) 22.7 | (660) 23.3 | (351) 26.2 | .13 |
| History of obstructive sleep apnea, (No.), % | (40) 20.4 | (126) 22.4 | (594) 22.1 | (636) 22.5 | (272) 20.3 | .56 |
| History of atrial fibrillation, (No.), % | (29) 14.8 | (75) 13.3 | (426) 15.8 | (471) 16.6 | (160) 11.9 | .001 |
| History of coronary artery disease, (No.), % | (45) 23.0 | (117) 20.8 | (697) 25.9 | (739) 26.1 | (274) 20.4 | <.001 |
| History of vascular disease, (No.), % | (13) 6.6 | (46) 8.2 | (243) 9.0 | (255) 9.0 | (105) 7.8 | .51 |
| History of major bleed, (No.), % | (50) 25.5 | (126) 22.4 | (657) 24.4 | (726) 25.7 | (298) 22.2 | .12 |
| History of congestive heart failure, (No.), % | (37) 18.9 | (91) 16.2 | (529) 19.7 | (601) 21.2 | (255) 19.0 | .06 |
| History of asthma, (No.), % | (60) 30.6 | (203) 36.1 | (950) 35.3 | (1032) 36.5 | (500) 37.2 | .38 |
| History of COPD, (No.), % | (40) 20.4 | (132) 23.4 | (658) 24.5 | (738) 26.1 | (350) 26.1 | .21 |
| History of pulmonary embolism, (No.), % | (14) 7.1 | (16) 2.8 | (119) 4.4 | (140) 4.9 | (53) 3.9 | .06 |
| History of chronic kidney disease, (No.), % | (43) 21.9 | (88) 15.6 | (497) 18.5 | (594) 21.0 | (208) 15.5 | <.001 |
| History of cancer, (No.), % | (40) 20.4 | (94) 16.7 | (548) 20.4 | (588) 20.8 | (200) 14.9 | <.001 |
| History of hypothyroidism, (No.), % | (38) 19.4 | (96) 17.1 | (567) 21.1 | (607) 21.5 | (234) 17.4 | .008 |
| History of low back pain, (No.), % | (63) 32.1 | (202) 35.9 | (958) 35.6 | (1000) 35.3 | (439) 32.7 | .34 |
| History of ascites, (No.), % | (50) 25.5 | (115) 20.4 | (719) 26.7 | (723) 25.6 | (277) 20.7 | <.001 |
| History of anxiety, (No.), % | (35) 17.9 | (96) 17.1 | (544) 20.2 | (557) 19.7 | (304) 22.7 | .05 |
| History of depression, (No.), % | (35) 17.9 | (107) 19.0 | (499) 18.6 | (507) 17.9 | (264) 19.7 | .73 |
| CCI total score, mean (median) | 1.8, 1 | 1.5, 1 | 1.7, 1 | 1.7, 1 | 1.5, 1 | <.001 |
| Estimated risk (%) of mortality within 90 days after hospitalization, mean (median) | 29.0, 22 | 22.8, 16 | 27.8, 24 | 26.9, 22 | 20.9, 14 | <.001 |
| **Medications** |  |  |  |  |  |  |
| ACE, (No.), % | (40) 20.4 | (128) 22.7 | (661) 24.6 | (670) 23.7 | (305) 22.7 | .51 |
| ARB, (No.), % | (22) 11.2 | (88) 15.6 | (422) 15.7 | (430) 15.2 | (183) 13.6 | .25 |
| Beta-blocker, (No.), % | (91) 46.4 | (220) 39.1 | (1213) 45.1 | (1279) 45.2 | (506) 37.7 | <.001 |
| Calcium channel blocker, (No.), % | (53) 27.0 | (140) 24.9 | (786) 29.2 | (798) 28.2 | (363) 27.1 | .24 |
| Diuretic, (No.), % | (70) 35.7 | (190) 33.7 | (1022) 38.0 | (1062) 37.5 | (430) 32.1 | .002 |
| DOACS, (No.), % | (33) 16.8 | (66) 11.7 | (399) 14.8 | (411) 14.5 | (147) 11.0 | .003 |
| Warfarin, (No.), % | (13) 6.6 | (29) 5.2 | (177) 6.6 | (184) 6.5 | (49) 3.7 | .002 |
| Anti-platelet, (No.), % | (68) 34.7 | (186) 33.0 | (980) 36.5 | (1007) 35.6 | (375) 28.0 | <.001 |
| Insulin, (No.), % | (43) 21.9 | (115) 20.4 | (504) 18.7 | (499) 17.6 | (205) 15.3 | .02 |
| Metformin, (No.), % | (33) 16.8 | (113) 20.1 | (544) 20.2 | (525) 18.6 | (224) 16.7 | .07 |
| Statin, (No.), % | (109) 55.6 | (295) 52.4 | (1519) 56.5 | (1601) 56.6 | (582) 43.4 | <.001 |
| Short acting bronchodilator, (No.), % | (52) 26.5 | (138) 24.5 | (689) 25.6 | (792) 28.0 | (350) 26.1 | .24 |
| Anti-depressant, (No.), % | (89) 45.4 | (204) 36.2 | (1034) 38.5 | (1102) 39.0 | (459) 34.2 | .005 |
| Corticosteroids, (No.), % | (67) 34.2 | (216) 38.4 | (1097) 40.8 | (1125) 39.8 | (528) 39.4 | .37 |
| Prednisone, (No.), % | (21) 10.7 | (74) 13.1 | (353) 13.1 | (367) 13.0 | (175) 13.1 | .92 |
| Immunomodulators, (No.), % | (4) 2.0 | (7) 1.2 | (62) 2.3 | (71) 2.5 | (23) 1.7 | .26 |
| Muscle relaxant, (No.), % | (19) 9.7 | (64) 11.4 | (316) 11.8 | (300) 10.6 | (168) 12.5 | .36 |
| NSAIDS, (No.), % | (56) 28.6 | (163) 29.0 | (720) 26.8 | (758) 26.8 | (382) 28.5 | .63 |
| Opioids, (No.), % | (42) 21.4 | (147) 26.1 | (709) 26.4 | (757) 26.8 | (314) 23.4 | .10 |
| **Laboratory values** |  |  |  |  |  |  |
| White blood cells, mean, median | 7.6, 6 | 7.3, 6 | 8.1, 7 | 8.6, 7 | 7.6, 7 | <.001 |
| Lymphocytes, mean, median | 15.5, 13 | 16.4, 14 | 14.7, 13 | 13.8, 12 | 15.8, 13 | <.001 |
| Neutrophils, mean, median | 74.6, 76 | 73.1, 75 | 75.1, 78 | 76.2, 78 | 74.3, 77 | <.001 |
| Hemoglobin, mean, median | 12.4, 13 | 12.7, 13 | 12.7, 13 | 12.7, 13 | 13.0, 13 | <.001 |
| Platelets, mean, median | 218.9, 195 | 214.6, 198 | 210.0, 195 | 221.4, 206 | 216.2, 200 | <.001 |
| AST, mean, median | 56.1, 38 | 45.8, 31 | 55.6, 34 | 57.2, 34 | 51.5, 35 | <.001 |
| ALT, mean, median | 42.4, 28 | 37.2, 27 | 42.4, 27 | 45.1, 28 | 42.4, 30 | <.001 |
| Creatinine, mean, median | 1.6, 1 | 1.4, 1 | 1.5, 1 | 1.6, 1 | 1.5, 1 | <.001 |
| Neutrophil to lymphocyte ratio, (No.), % |  |  |  |  |  |  |
| Less than or equal to 3.0 | (56) 21.5 | (150) 22.5 | (633) 19.3 | (568) 16.7 | (360) 21.5 | <.001 |
| 3.0 to 6.0 | (80) 30.7 | (228) 34.2 | (977) 29.7 | (950) 28.0 | (521) 31.1 |  |
| More than 6.0 | (125) 47.9 | (288) 43.2 | (1676) 51.0 | (1876) 55.3 | (795) 47.4 |  |
| Systemic Inflammatory Index, (No.), % |  |  |  |  |  |  |
| SII <=300 | (17) 6.6 | (64) 9.6 | (251) 7.7 | (213) 6.3 | (133) 8.0 | <0.001 |
| SII >300 to <=600 | (48) 18.5 | (133) 20.0 | (558) 17.0 | (457) 13.5 | (293) 17.5 |  |
| SII >600 to <=900 | (40) 15.4 | (103) 15.5 | (477) 14.5 | (456) 13.4 | (288) 17.2 |  |
| SII > 900 | (154) 59.5 | (366) 55.0 | (1997) 60.8 | (2266) 66.8 | (960) 57.3 |  |

Wave 1: - March 19 – June 16, 2020; Wave 2: June 17 – September 19, 2020; Wave 3a: September 20 December 13, 2020; Wave 3b: December 14, 2020 – March 10, 2021; Wave 4: March 11 – June 6, 2021.

## References

1. Gates B. Innovation for pandemics. New England Journal of Medicine. 2018 May 31;378(22):2057–2060. doi: 10.1056/NEJMp1806283. PMID: 29847763.

2. Gates B. Responding to Covid-19 - A once-in-a-century pandemic? New England Journal of Medicine. 2020 Apr 30;382(18):1677–1679. doi: 10.1056/NEJMp2003762. Epub 2020 Feb 28. PMID: 32109012.

3. Hechenbleikner EM, Samarov DV, Lin E. Data explosion during COVID-19: A call for collaboration with the tech industry & data scrutiny. EClinicalMedicine. 2020 May 24;23:100377. doi: 10.1016/j.eclinm.2020.100377. PMID: 32632412; PMCID: PMC7245577.

4. Romanelli RJ, Azar KMJ, Sudat S, Hung D, Frosch DL, Pressman AR. Learning health system in crisis: Lessons from the COVID-19 pandemic. Mayo Clin Proc Innov Qual Outcomes. 2021 Feb;5(1):171–176. doi: 10.1016/j.mayocpiqo.2020.10.004. Epub 2020 Oct 29. PMID: 33163894; PMCID: PMC7598312.

5. Institute of Medicine (US) Roundtable on Evidence-Based Medicine. The Learning Healthcare System: Workshop Summary. Olsen L, Aisner D, McGinnis JM, editors. Washington (DC): National Academies Press (US); 2007. PMID: 21452449.

6. UPMC REMAP-COVID Group, on behalf of the REMAP-CAP Investigators. Implementation of the Randomized Embedded Multifactorial Adaptive Platform for COVID-19 (REMAP-COVID) trial in a US health system-lessons learned and recommendations. Trials. 2021 Jan 28;22(1):100. doi: 10.1186/s13063-020-04997-6. Erratum in: Trials. 2021 Feb 16;22(1):145. PMID: 33509275; PMCID: PMC7841377.

7. Huang DT, McCreary EK, Bariola JR, Wadas RJ, Kip KE, Marroquin OC, Koscumb S, Collins K, Shovel JA, Schmidhofer M, Wisniewski MK, Sullivan C, Yealy DM, Axe M, Nace DA, Haidar G, Khadem T, Linstrum K, Snyder GM, Seymour CW, Montgomery SK, McVerry BJ, Berry L, Berry S, Meyers R, Weissman A, Peck-Palmer OM, Wells A, Bart R, Albin DL, Minnier T, Angus DC. The UPMC OPTIMISE-C19 (OPtimizing Treatment and Impact of Monoclonal antIbodieS through Evaluation for COVID-19) trial: a structured summary of a study protocol for an open-label, pragmatic, comparative effectiveness platform trial with response-adaptive randomization. Trials. 2021 May 25;22(1):363. doi: 10.1186/s13063-021-05316-3. PMID: 34034784; PMCID: PMC8144687.

8. Benchimol EI, Smeeth L, Guttmann A, Harron K, Moher D, Petersen I, Sørensen HT, von Elm E, Langan SM; RECORD Working Committee. The REporting of studies Conducted using Observational Routinely-collected health Data (RECORD) statement. PLoS Med. 2015 Oct 6;12(10):e1001885. doi: 10.1371/journal.pmed.1001885. PMID: 26440803; PMCID: PMC4595218.

9. Horby P, Kim WS, Emberson J, Mafham M, Bell J, Linsell L, … RECOVERY Collaborative Group. Effect of dexamethasone in hospitalized patients with COVID-19 – preliminary report. MedRxiv 2020.06.22.20137273 doi: https://doi.org/10.1101/2020.06.22.20137273

10. RECOVERY Collaborative Group, Horby P, Lim WS, Emberson JR, Mafham M, Bell JL, Linsell L, Staplin N, Brightling C, Ustianowski A, Elmahi E, Prudon B, Green C, Felton T, Chadwick D, Rege K, Fegan C, Chappell LC, Faust SN, Jaki T, Jeffery K, Montgomery A, Rowan K, Juszczak E, Baillie JK, Haynes R, Landray MJ. Dexamethasone in hospitalized patients with Covid-19. New England Journal of Medicine. 2021 Feb 25;384(8):693–704. doi: 10.1056/NEJMoa2021436. Epub 2020 Jul 17. PMID: 32678530; PMCID: PMC7383595.

11. Food and Drug Administration (FDA). Coronavirus (COVID-19) Update: FDA Issues Emergency Use Authorization for Potential COVID-19 Treatment. https://www.fda.gov/news-events/press-announcements/coronavirus-covid-19-update-fda-issues-emergency-use-authorization-potential-covid-19-treatment. May 1, 2020. Accessed June 10, 2021.

12. Gilead Press Releases. Gilead Announces Results from Phase 3 Trial of Remdesivir in Patients with Moderate COVID-19. https://www.gilead.com/news-and-press/press-room/press-releases/2020/6/gilead-announces-results-from-phase-3-trial-of-remdesivir-in-patients-with-moderate-covid-19. June 1, 2020. Accessed June 10, 2021.

13. U.S. Health and Human Services. Trump Administration Secures New Supplies of Remdesivir for the United States. https://public3.pagefreezer.com/browse/HHS%20%E2%80%93%C2%A0About%20News/20-01-2021T12:29/ https://www.hhs.gov/about/news/2020/06/29/trump-administration-secures-new-supplies-remdesivir-united-states.html. June 29, 2020. Accessed June 10, 2021.

14. U.S. Food and Drug Administration (FDA). FDA Approves First Treatment for COVID-19. https://www.fda.gov/news-events/press-announcements/fda-approves-first-treatment-covid-19#:∼:text=Today%2C%20the%20U.S.%20Food%20and,of%20COVID%2D19%20requiring%20hospitalization. October 22, 2020. Accessed June 10, 2021

15. U.S. Food and Drug Administration (FDA). Request for Emergency Use Authorization for Use of Chloroquine Phosphate or Hydroxychloroquine Sulfate Supplied from the Strategic National Stockpile for Treatment of 2019 Coronavirus Disease. https://www.fda.gov/media/136534/download. March 28, 2020. Accessed June 10, 2021.

16. Solender A. All the Times Trump Has Promoted Hydroxychloroquine. Forbes Magazine, May 22, 2020. https://www.forbes.com/sites/andrewsolender/2020/05/22/all-the-times-trump-promoted-hydroxychloroquine/?sh=6c6ba53b4643. Accessed June 10, 2021.

17. U.S. Food and Drug Administration (FDA). FDA cautions against use of hydroxychloroquine or chloroquine for COVID-19 outside of the hospital setting or a clinical trial due to risk of heart rhythm problems. https://www.fda.gov/media/137250/download. April 24, 2020. Accessed June 10, 2021.

18. U.S. Food and Drug Administration (FDA). Memorandum Explaining Basis for Revocation of Emergency Use Authorization for Emergency Use of Chloroquine Phosphate and Hydroxychloroquine Sulfate. https://www.fda.gov/media/138945/download. June 15, 2020. Accessed June 10, 2021.

19. The REMAP-CAP Investigators, Gordon AC, Mouncey PR, Al-Beidh F, Rowan KM, Nichol AD, Arabi YM, et al. Interleukin-6 receptor antagonists in critically ill patients with Covid-19 – Preliminary report. MedRxiv 2021.01.09.2021. https://doi.org/10.1101/2021.01.07.21249390. January 9, 2021

20. Horby PW, Pessoa-Amorim G, Peto L, Brightling CE, Sarkar R, Thomas K, et al.: RECOVERY Collaborative Group. Tocilizumab in patients admitted to hospital with COVID-19 (RECOVERY): preliminary results of a randomised, controlled, open label platform trial. MedRxiv 2021.02.11.21249528: doi: https://doi.org/10.1101/2021.02.11.21249258

21. REMAP-CAP Investigators, Gordon AC, Mouncey PR, Al-Beidh F, Rowan KM, Nichol AD, Arabi YM, et al. Interleukin-6 receptor antagonists in critically ill patients with Covid-19. New England Journal of Medicine. 2021 Apr 22;384(16):1491–1502. doi: 10.1056/NEJMoa2100433. Epub 2021 Feb 25. PMID: 33631065; PMCID: PMC7953461.

22. Institute of Medicine (US) Roundtable on Evidence-Based Medicine. Leadership Commitments to Improve Value in Healthcare: Finding Common Ground: Workshop Summary. Washington (DC): National Academies Press (US); 2009. PMID: 21391347.

23. McGinnis JM, Fineberg HV, Dzau VJ. Advancing the Learning Health System. New England Journal of Medicine. 2021 Jul 1;385(1):1–5. doi: 10.1056/NEJMp2103872. Epub 2021 Jun 26. PMID: 34192452.

24. Kirkham JJ, Penfold NC, Murphy F, Boutron I, Ioannidis JP, Polka J, Moher D. Systematic examination of preprint platforms for use in the medical and biomedical sciences setting. BMJ Open. 2020 Dec 29;10(12):e041849. doi: 10.1136/bmjopen-2020-041849. PMID: 33376175; PMCID: PMC7778769.

25. Bauchner H, Fontanarosa PB, Golub RM. Editorial evaluation and peer review during a pandemic: How journals maintain standards. JAMA. 2020 Aug 4;324(5):453–454. doi: 10.1001/jama.2020.11764. PMID: 32589195.

26. Park JJH, Mogg R, Smith GE, Nakimuli-Mpungu E, Jehan F, Rayner CR, Condo J, Decloedt EH, Nachega JB, Reis G, Mills EJ. How COVID-19 has fundamentally changed clinical research in global health. Lancet Glob Health. 2021 May;9(5):e711–e720. doi: 10.1016/S2214-109X(20)30542-8. PMID: 33865476; PMCID: PMC8049590.

27. Alexander PE, Debono VB, Mammen MJ, Iorio A, Aryal K, Deng D, Brocard E, Alhazzani W. COVID-19 coronavirus research has overall low methodological quality thus far: case in point for chloroquine/hydroxychloroquine. J Clin Epidemiol. 2020 Jul;123:120–126. doi: 10.1016/j.jclinepi.2020.04.016. Epub 2020 Apr 21. PMID: 32330521; PMCID: PMC7194626.

28. Morris ZS, Wooding S, Grant J. The answer is 17 years, what is the question: understanding time lags in translational research. J R Soc Med. 2011 Dec;104(12):510–20. doi: 10.1258/jrsm.2011.110180. PMID: 22179294; PMCID: PMC3241518.

29. Melnyk BM, Fineout-Overholt E, Gallagher-Ford L, Kaplan L. The state of evidence-based practice in US nurses: critical implications for nurse leaders and educators. Journal of Nursing Administration. 2012 Sep;42(9):410–7. doi: 10.1097/NNA.0b013e3182664e0a. PMID: 22922750.

30. Warren JI, McLaughlin M, Bardsley J, Eich J, Esche CA, Kropkowski L, Risch S. The strengths and challenges of implementing EBP in healthcare systems. Worldviews Evid Based Nurs. 2016 Feb;13(1):15–24. doi: 10.1111/wvn.12149. PMID: 26873372.

31. Saville BR, Berry SM. Balanced covariates with response adaptive randomization. Pharm Stat. 2017 May;16(3):210–217. doi: 10.1002/pst.1803. Epub 2017 Mar 6. PMID: 28261972.

32. Angus DC, Derde L, Al-Beidh F, Annane D, Arabi Y, Beane A, et al. Effect of hydrocortisone on mortality and organ support in patients with severe COVID-19: The REMAP-CAP COVID-19 Corticosteroid Domain Randomized Clinical Trial. JAMA. 2020 Oct 6;324(13):1317–1329. doi: 10.1001/jama.2020.17022. PMID: 32876697; PMCID: PMC7489418.

33. Beck AF, Hartley DM, Kahn RS, Taylor SC, Bishop E, Rich K, Saeed MS, Schuler CL, Seid M, Cronin SC, Raney L, Zafar MA, Margolis PA. Rapid, bottom-up design of a regional learning health system in response to COVID-19. Mayo Clin Proc. 2021 Apr;96(4):849–855. doi: 10.1016/j.mayocp.2021.02.006. Epub 2021 Feb 16. PMID: 33714596; PMCID: PMC7885665.

34. Khanna N, Klyushnenkova EN, Kaysin A, Stewart DL. Utilizing the learning health system adaptation to guide family medicine practice to COVID-19 response. J Prim Care Community Health. 2020 Jan-Dec;11:2150132720966409. doi: 10.1177/2150132720966409. PMID: 33063617; PMCID: PMC7580135.

35. Anesi GL, Jablonski J, Harhay MO, Atkins JH, Bajaj J, Baston C, et al. Characteristics, outcomes, and trends of patients with COVID-19-related critical illness at a learning health system in the United States. Ann Intern Med. 2021 May;174(5):613–621. doi: 10.7326/M20-5327. Epub 2021 Jan 19. PMID: 33460330; PMCID: PMC7901669.

